# Burden of cardiometabolic risk factors and preclinical target organ damage among adults in Freetown, Sierra Leone: a community-based health-screening survey

**DOI:** 10.1101/2023.02.20.23286145

**Authors:** James Baligeh Walter Russell, Theresa Ruba Koroma, Santigie Sesay, Sallieu K Samura, Sulaiman Lakoh, Ansu Bockarie, Onomeh Thomas Abiri, Joseph Sam Kanu, Joshua Coker, Abdul Jalloh, Victor Conteh, Sorie Conteh, Mohamed Smith, Othman. Z. Mahdi, Durodami. R. Lisk

## Abstract

**Objective:** The aim of the study was to investigate the prevalence of cardiometabolic risk factors (CMRFs), target organ damage and its associated factors among adults in Freetown, Sierra Leone.

**Design:** This community-based cross-sectional study used a stratified multistage random sampling method to recruit adult participants.

**Setting:** The health screening study was conducted between October 2019 and October 2021 in Western Area Urban, Freetown, Sierra Leone.

**Participants:** A total of 2394 adults Sierra Leoneans aged 20 years, or more were enrolled.

**Outcome measure:** Anthropometric data, fasting lipid profiles, fasting plasma glucose, target organ damage, clinical profiles and demographic characteristic of participants were described. The cardiometabolic risks were further related to target organ damage.

**Results:** The prevalence of known CMRFs was 35.3% for hypertension, 8.3% for diabetes mellitus, 21.1% for dyslipidemia, 10.0% for obesity, 13.4% for smoking and 37.9% for alcohol. Additionally, 16.1% had left ventricular hypertrophy (LVH) by electrocardiogram (ECG), 14.2% had LVH by 2D-Echo, and 11.4% had chronic kidney disease. The odds of developing ECG-LVH were higher with diabetes [OR = 1.255, 95% C.I. (0.822 - 1.916) and dyslipidaemia [OR = 1.449, 95% C.I. (0.834 - 2.518). The association factors for higher odds of LVMI by echo were dyslipidemia [OR = 1.844, 95% C.I (1.006-3.380)] and diabetes mellitus [OR =1.176, 95% C.I. (0.759-1.823)]. The odds of having CKD were associated with diabetes mellitus [OR =1.212, 95% CI (0.741-1.983)] and hypertension [OR =1.163, 95% CI (0.887-1.525)]. A low optimal cut-off point for ECG-LVH (male 24.5mm vs female 27.5mm) as a target organ damage was required to maximize sensitivity and specificity by a receiver operating characteristic (ROC) curve since the odds for LVH by ECG was low.

**Conclusions:** This study provides novel data-driven information on the burden of cardiometabolic risks and its association with preclinical target organ damage in a resource limited setting. It illustrates the need for interventions in improve cardiometabolic health screening and management among adults in Sierra Leoneans.

**Strengths and Limitations of the study:** 

**Strengths:**

- A major strength of this study is its community-based design and the first study of its kind on a larger population in Sierra Leone.
- The study was statistically powered to produce results that are representative of adults in Sierra Leone.

**Limitations:**

- The study is limited as it could not conclude direct causality inference of risk factors and effect outcomes.
- Since some of the outcomes (fasting plasma glucose, HbA1c and fasting lipid profile) are limited by the reliance on single time point measurements, it may result in measurement errors and the potential of underestimating cardiometabolic risk factors.
- Chronic kidney disease (CKD) assessment by single serum creatinine without assessing for proteinuria, which also indicates the presence of CKD, will lead to underestimation of CKD.

## INTRODUCTION

Cardiometabolic diseases (CMD) are a group of complex disorders, including cardiovascular diseases and diabetes mellitus. The spectrum of cardiometabolic disease begins with insulin resistance, a trait that is expressed early in life and later will progress to clinically identifiable high-risk states of prediabetes, then to type 2 diabetes mellitus (T2DM) and cardiovascular diseases (CVD)[1].

CVD is of great interest because its insidious progression is marked by a multistage pathogenesis that is often heralded by asymptomatic changes in the heart, kidney, and blood vessels [1, 2]. The associated risk factors of CMD are a cluster of obesity (particularly central adiposity), dyslipidaemia, psychosocial stress, and unhealthy lifestyles like physical inactivity, lack of consumption of fruits/vegetables, cigarette smoking, and harmful alcohol consumption [2, 3]. These risk factors are associated with dysfunctional biomedical processes within the body, with the potential of triggering cardiovascular diseases (CVD) and their related complications of chronic non-communicable diseases (NCDs) [4-6]

According to World Health Organization (WHO), NCDs are the leading causes of morbidity and mortality, with more than three-quarters of NCDs deaths occurring in low-and-middle-income countries [3]. In 2017, the Global Burden of Disease Study reported a dramatic increase in the total number of deaths in NCD by 22.7% (21.5% -23.9%) from 2007 to 2017, while the Disability-adjusted-life years (DALYs) related to CVDs was 73.3%. During the same period (2007 – 2017), there was an estimated increase of 7.61 million deaths, with the highest rate in western sub-Saharan Africa (SSA) [7]. This epidemiological transition from communicable to non-communicable diseases in SSA has resulted in an exponential rise in cardiometabolic risk factors (CMRF) [8]. The recent demographic transition witnessed in urban settings of many LMICs may be attributed to adopting western lifestyle behaviours, including, poor eating habits, harmful alcohol consumption and cigarette smoking [9-11]. These settings will also be a favourable platform for developing cardiometabolic risk factors and its attending target organ damage. While there is recognition of the rising burden of NCDs across Africa, scanty information exists in most SSA countries because of the absence of well-developed health programmes for the comprehensive evaluation and management of high-risk individuals [12, 13]. Our understanding of this spectrum of diseases is disproportionately informed by studies conducted in developed countries. Such findings may not be entirely applicable to individuals in developing countries. Reasons for this could be related to differences in genetic characteristics and CVDs risk factors across countries and regions[14].

Sierra Leone is one of the least developed countries in the world, with a double burden of communicable and non-communicable diseases. The eleven years of devastating civil war (1991 – 2002) disrupted the health system, and its long-term effects were still seen during the public health crisis caused by the 2013-2016 Ebola outbreak [15, 16]. Since the civil war, Sierra Leone has experienced significant urbanization in recent years, and this demographic evolution has impacted on the socioeconomic growth recovery of the nation. This type of chaotic urbanization also referred to as a “complex urban health crisis” is seen in other SSA countries because it serves as a harbinger that is accelerating non-communicable disease burden and is an existential threat to the health and development of a nation [17-19].

In Sierra Leone, the evaluation of CVDs has been conducted by several small studies but with very little information on the assessment of CMRF burden [20-23]. Although a recent survey in a provincial district setting (rural and urban) suggested a high prevalence of CMRF, there is limited data estimating preclinical target organ damage in this West African country [24]. Additionally, there is no report of a direct evaluation of CMRF in any settlement in the capital city of Sierra Leone. This study aimed to comprehensively evaluate the prevalence of cardiovascular risk factors and TOD in a population-based study in Freetown, Sierra Leone. The study also investigated how these known CVDRFs are associated with the preclinical cardiac and renal TOD among adults aged 20 years or more.

## METHODS

### Patient and public involvement

Patients and/or the public were not involved in the design, or conduct, or reporting, or dissemination plans of this research.

### Study setting and design

This population-based cross-sectional study was a health screening survey conducted between October 2019 and October 2021 among adults living in Western Area Urban, Freetown, Sierra Leone. It was a screening and awareness programme for non-communicable diseases in Western Area Urban, initiated and funded by Ecobank Sierra Leone Limited. Freetown is the capital city of Sierra Leone, with an estimated 1.5 million inhabitants [25]. Freetown is important because of its densely heterogeneous population and the main business centre of Sierra Leone. It sets the trend for the rest of the country as its demographic distribution is similar to other larger cities. All ethnic groups in the country can be found in Freetown, with Krio and English being the primary spoken languages.

### Sample size calculation, participant recruitment, and selection

The study was designed to provide results that truly represent the adult population in Sierra Leone. A month before the awareness and screening campaign for non-communicable diseases, citizens within the Freetown municipality were informed about these activities by repeated mass communication through National Radio and Television stations. We used a stratified random sampling strategy to recruit adult Sierra Leonean participants aged ≥ 20. Western Area Urban - Freetown is divided into eight official electoral constituencies (Central I, II, East I, II, III, & West I, II, III), and the first stage in the sampling strategy was to select all eight constituencies. This was followed by subdividing each constituent region into subzones using the 2015 census data [25], and subsequently, one of the sub-zonal communities was selected by simple random sampling. The selected communities were namely: Calabar Town - East III, Low-Cost Housing Community - East II, Ginger Hall Community - East I, Mountain Cut Community - Central II, PWD/Pademba Road community - Central I, Brookfield’s Community – West II, and Aberdeen Community – West III. Potential participants within each selected sub-zonal community were line-listed at their community health centre, and these enlisted individuals were selected by simple random sampling methods. The following participants were excluded: Pregnant and lactating mothers, those with mental illness/dementia and persons unwilling to grant consent. The sample size was calculated using the clinical estimated prevalence of 22% for hypertension in Sierra Leone [26]. The minimum sample size was assessed using the Leisle Kish formula [27]:

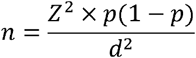

where *n* is the sample size (number of adult participants), *p* is the expected prevalence of hypertension in an adult population (*p* = 0.22), and *d* is the precision (if 5%, *d* = 0.05). The Z value is 1.96 for a 95% Confidence Interval (C.I).

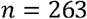

To minimize bias and allow attrition of non-response and non-availability of data, the sample size was oversampled by 20%.

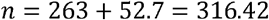

Using a design effect of eight sub-zonal communities to adjust the sample size:

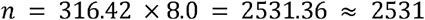

### Procedure and data collection

All eligible potential participants in each selected community were invited to participate in the ‘awareness and screening campaign for non-communicable disease’ on a designated date at the National Victoria Park. The WHO stepwise approach guided the process of data collection for this study. Medical students, doctors and nurses were trained on the campaign’s conduct, including data collection. (Flow chart during the campaign is shown in Fig:1).

### Demographic and health history

A standard questionnaire was used to obtain information on demographics (age, sex, and education), lifestyle (fruits and vegetable consumption, smoking status and physical activity) and medical history (family history of hypertension and diabetes mellitus). Translators were used for participants who could not understand English. An OMRON M3 electronic sphygmomanometer with an appropriate cuff size was used to record a participant’s blood pressure in the sitting position, and measurements were taken after at least 3-5 minutes of rest. The mean of the two recorded readings was taken as the participant’s blood pressure. Body weight, height, and waist circumference were measured with light clothes and bare feet.

### Outcome measures and definition

i. Hypertension was defined as an average SBP of 140 mm Hg or higher, or DBP of 90 mm Hg or greater or a participant reported current use of antihypertensive medication [28].
ii. A participant who smoked more than 100 sticks of cigarettes in his or her lifetime and still smoking at the interview was referred a smoker, while an ex-smoker was someone who had stopped smoking at least 28 days before the interview [29].
iii. Data on alcohol consumption was based on the WHO step survey tool [30].
iv. Physical activity was classified into “Low”, “Moderate”, and “Vigorous”.
  ○ Low physical activity: Sedentary lifestyles at work and home
  ○ Moderate physical activity: Brisk walking, domestic house chores and general house task such as roofing and painting, moderate farm work like weeding Vigorous physical activity: running, briskly ascending and descending hill tasks, intense farm working and carrying masses > 20kg [31]
v. The body mass index (BMI) was calculated as a ratio of the weight in kilograms and the square of the height in metres. BMI-based body habitus (in kg/m^2^) was classified as underweight (BMI <18.5), normal weight (BMI=18.5–24.9), overweight (BMI=25.0–29.9) and obese (BMI ≥30) [32].

### Cardiometabolic risk factors definition

The cardiometabolic risk factors measured in this study include blood pressure, fasting blood sugar, HbA1c, waist circumference, BMI, and serum lipids.

vi Overall obesity was defined as BMI ≥ 30 kg/m^2^
vii Abdominal obesity was defined as waist circumference > 88 cm for women and 102 cm for men [33].
viii Diabetes mellitus was defined as a fasting blood glucose (FBG) level of 7.0 mmol/L or greater, HbA1c ≥ 6.5%, or the use of insulin or an oral hypoglycaemic agent. Pre-diabetes was defined as FPG between 6.1mmol/l (110 mg/ dL) and 6.9 mmol/l (124.9 mg/dL) [34].

At the health screening venue (Victoria Park), consented and enrolled participants who had completed their screening questionnaires were referred for cardiac evaluation, ECG, echocardiographic and to an accredited reference laboratory for blood sample collection.

### Clinical biochemistry measurements

Participants’ blood samples were collected from the median cubital vein between 8:00 and 10:00 AM, after overnight fasting for 8 to 10 hours. These samples were processed within 4 hours of collection per manufacturers’ instructional protocols, using Beckman Coulter: AU480 Chemistry System. Glucose, total cholesterol, triglycerides, high-density lipoprotein (HDL-C) and low-density lipoprotein (LDL-C) were analysed. America Diabetes Association cut points were used to evaluate lipid panel markers and DM abnormalities. Dyslipidaemia was defined as TG ≥ 1.70 mmol/L (150 mg/dL), TC ≥ 6.22 mmol/L (240 mg/dL), LDL ≥ 3.3mmol/L (130 mg/dL), HDL <1.04 mmol/L (40 mg/dL), use of lipid-lowering medications, was considered an abnormal high [35].

### Preclinical TOD definition

A cardiologist evaluated each participant for cardiac target organ damage using transthoracic echocardiography (GE vivid e ultrasound system equipped with MSR-RS 1.5 to 3.5 MHz sector and linear probe). The recommended formula for calculating left ventricular mass was used by measuring the 2-dimensional guided M-mode imaging. The LVM index was calculated by dividing LVM by body surface area (See Data Supplemental Method 1). Cardiac TOD for Left ventricular hypertrophy (LVH) was defined as left ventricular mass index (LVMI) > 95 g/m^2^ for women and > 115 g/m^2^ for men, according to the American Society of Echocardiography (ASE) recommendation [36]. Renal TOD was evaluated by using the estimated glomerular filtration rate (eGFR), an essential chronic kidney disease (CKD) marker [37] (Data Supplemental Method 2).

### Statistical Analysis

Data analysis was done using IBM SPSS Statistical 2.6 and STATA 17 software. Baseline characteristics, cardiometabolic risk factors and target organ damage characteristics were analyzed by sex and zones. Categorical variables were expressed as numbers and percentages, and the Pearson chi-square (X^2^) test was used to assess the difference. Continuous variables were expressed as mean ± SD and compared using a one-way analysis of variance (ANOVA). Median and IQR were used when necessary. Multivariable logistic regression was done to determine associations between demographic characteristics and cardiovascular risk factors. A two-tailed p-value of ≤ 0.05 was considered statistically significant. Subsequently, receiver operating characteristics (ROC) were conducted to evaluate and compare the sensitivity of the different cardiometabolic risk factors. A multivariate binary logistic regression with a forced entry for all independent variables was used to assess the odds of targeted organ damage (LVH, LVMI and CKD) associated with cardiometabolic risk factors. To determine the influence of potential confounders on the association between cardiometabolic risk factors and targeted organ damage, the following models were generated: Model 1 adjusted age and sex; Model 2 adjusted for age, sex, marital status, education level, income, and occupation.

## RESULTS

### Basic characteristics of the study

A total of 2531 participants were recruited into the study, with a response rate of 94.6%. We excluded 54 participants who were absent on the “screening and awareness campaign” day, 53 who refused blood sampling by venous puncture, and 30 whose ECG and Echocardiographic data were missing. Finally, 2394 participants (52.2% female) with a mean age of 41.9 ± 12.3 years (p=0.550) were included in the analysis. Participants from the eight sub-zonal communities were equally selected without significant differences in population distribution (p=0.950). The baseline socio-demographic and clinical characteristics of all the participants are shown in Tables 1 & 2. While unemployment (38.8%) and being single (39.9%) were high among the study participants, we also noted that most of the participants were earning less than five hundred leones (< US$30) a month. According to WHO criteria, 91.1% of the study population consumed less than three servings of vegetables and fruits per week. Compared with women, more men were physically active (54.4% versus 45.6%) and consumed alcohol (51.5% versus 48.1%).

**Table 1.**
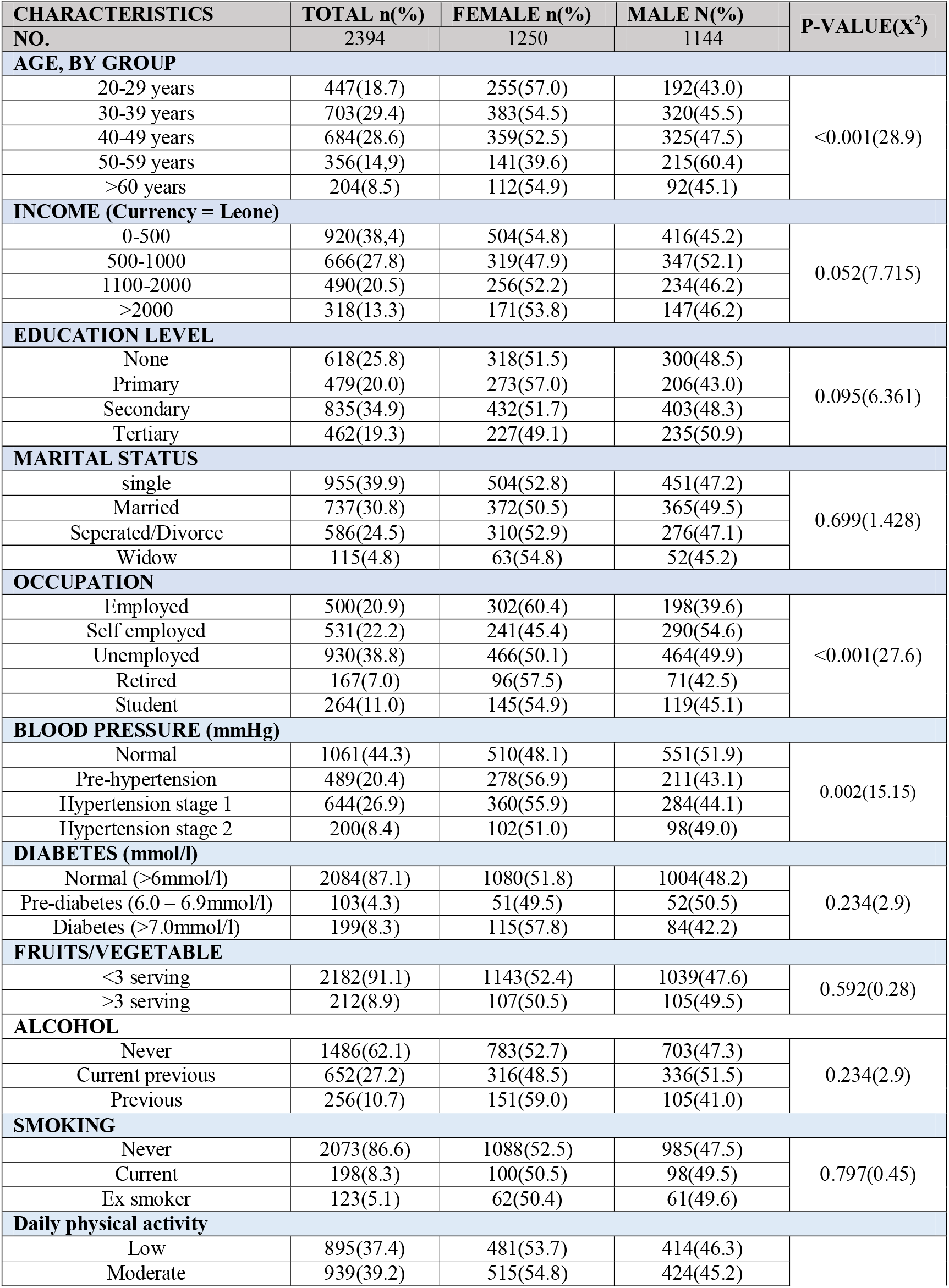
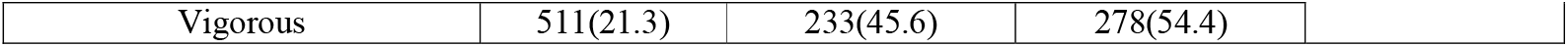
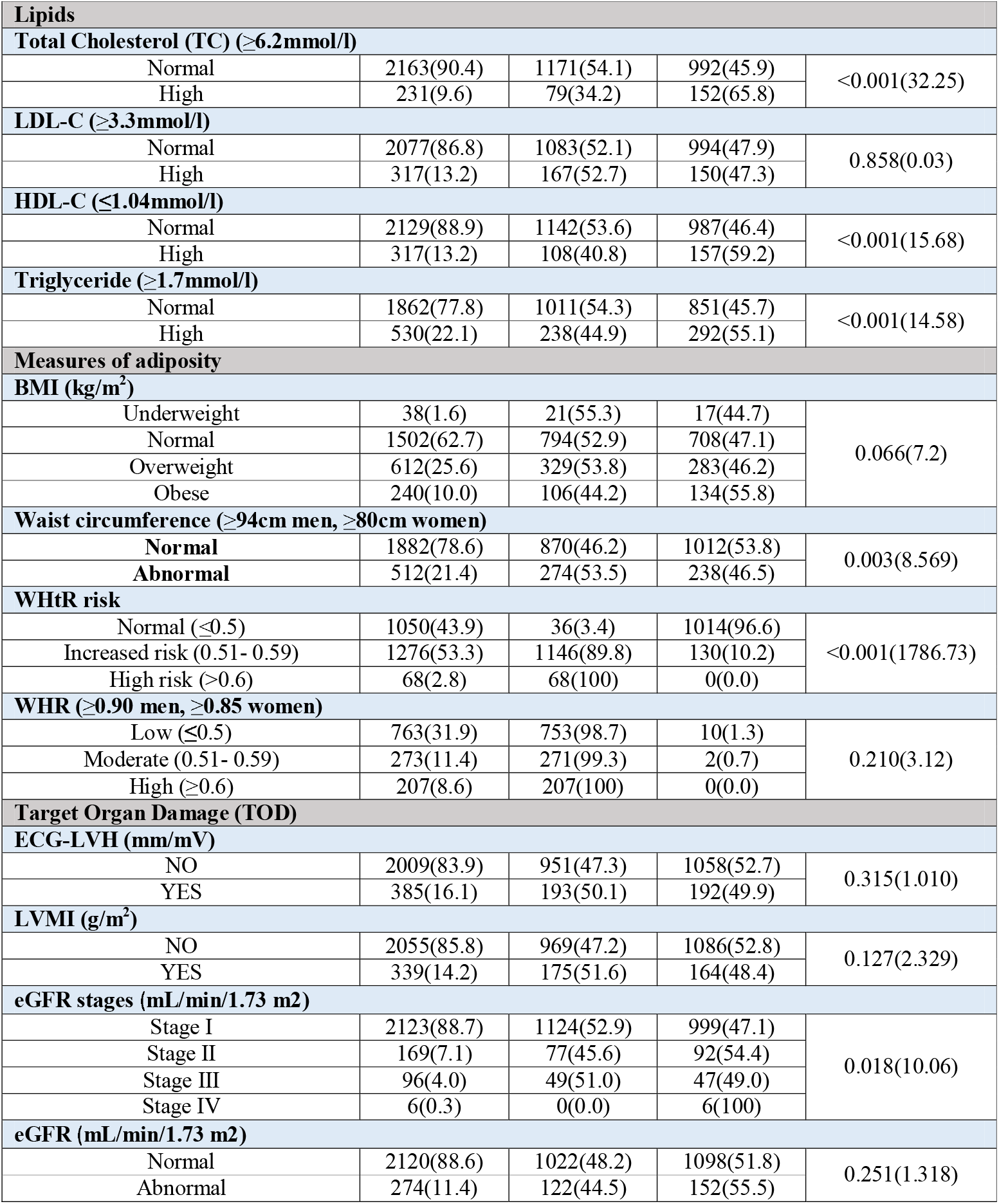
Socio-demographic and clinical characteristics of participants

| CHARACTERISTICS<br>NO. | TOTAL n(%) | FEMALE n(%) | MALE N(%) | P-VALUE(X<sup>2</sup>) |
| --- | --- | --- | --- | --- |
|  | 2394 | 1250 | 1144 |  |
| AGE, BY GROUP |  |  |  |  |
| 20-29 years | 447(18.7) | 255(57.0) | 192(43.0) | <0.001(28.9) |
| 30-39 years | 703(29.4) | 383(54.5) | 320(45.5) |  |
| 40-49 years | 684(28.6) | 359(52.5) | 325(47.5) |  |
| 50-59 years | 356(14.9) | 141(39.6) | 215(60.4) |  |
| >60 years | 204(8.5) | 112(54.9) | 92(45.1) |  |
| INCOME (Currency = Leone) |  |  |  |  |
| 0-500 | 920(38.4) | 504(54.8) | 416(45.2) | 0.052(7.715) |
| 500-1000 | 666(27.8) | 319(47.9) | 347(52.1) |  |
| 1100-2000 | 490(20.5) | 256(52.2) | 234(46.2) |  |
| >2000 | 318(13.3) | 171(53.8) | 147(46.2) |  |
| EDUCATION LEVEL |  |  |  |  |
| None | 618(25.8) | 318(51.5) | 300(48.5) | 0.095(6.361) |
| Primary | 479(20.0) | 273(57.0) | 206(43.0) |  |
| Secondary | 835(34.9) | 432(51.7) | 403(48.3) |  |
| Tertiary | 462(19.3) | 227(49.1) | 235(50.9) |  |
| MARITAL STATUS |  |  |  |  |
| single | 955(39.9) | 504(52.8) | 451(47.2) | 0.699(1.428) |
| Married | 737(30.8) | 372(50.5) | 365(49.5) |  |
| Seperated/Divorce | 586(24.5) | 310(52.9) | 276(47.1) |  |
| Widow | 115(4.8) | 63(54.8) | 52(45.2) |  |
| OCCUPATION |  |  |  |  |
| Employed | 500(20.9) | 302(60.4) | 198(39.6) | <0.001(27.6) |
| Self employed | 531(22.2) | 241(45.4) | 290(54.6) |  |
| Unemployed | 930(38.8) | 466(50.1) | 464(49.9) |  |
| Retired | 167(7.0) | 96(57.5) | 71(42.5) |  |
| Student | 264(11.0) | 145(54.9) | 119(45.1) |  |
| BLOOD PRESSURE (mmHg) |  |  |  |  |
| Normal | 1061(44.3) | 510(48.1) | 551(51.9) | 0.002(15.15) |
| Pre-hypertension | 489(20.4) | 278(56.9) | 211(43.1) |  |
| Hypertension stage 1 | 644(26.9) | 360(55.9) | 284(44.1) |  |
| Hypertension stage 2 | 200(8.4) | 102(51.0) | 98(49.0) |  |
| DIABETES (mmol/l) |  |  |  |  |
| Normal (>6mmol/l) | 2084(87.1) | 1080(51.8) | 1004(48.2) | 0.234(2.9) |
| Pre-diabetes (6.0 – 6.9mmol/l) | 103(4.3) | 51(49.5) | 52(50.5) |  |
| Diabetes (>7.0mmol/l) | 199(8.3) | 115(57.8) | 84(42.2) |  |
| FRUITS/VEGETABLE |  |  |  |  |
| <3 serving | 2182(91.1) | 1143(52.4) | 1039(47.6) | 0.592(0.28) |
| >3 serving | 212(8.9) | 107(50.5) | 105(49.5) |  |
| ALCOHOL |  |  |  |  |
| Never | 1486(62.1) | 783(52.7) | 703(47.3) | 0.234(2.9) |
| Current previous | 652(27.2) | 316(48.5) | 336(51.5) |  |
| Previous | 256(10.7) | 151(59.0) | 105(41.0) |  |
| SMOKING |  |  |  |  |
| Never | 2073(86.6) | 1088(52.5) | 985(47.5) | 0.797(0.45) |
| Current | 198(8.3) | 100(50.5) | 98(49.5) |  |
| Ex smoker | 123(5.1) | 62(50.4) | 61(49.6) |  |
| Daily physical activity |  |  |  |  |
| Low | 895(37.4) | 481(53.7) | 414(46.3) |  |
| Moderate | 939(39.2) | 515(54.8) | 424(45.2) |  |
| Vigorous | 511(21.3) | 233(45.6) | 278(54.4) |  |

**Table 1 Continued B**
| Lipids |  |  |  |  |
| --- | --- | --- | --- | --- |
| Total Cholesterol (TC) (≥6.2mmol/l) |  |  |  |  |
| Normal | 2163(90.4) | 1171(54.1) | 992(45.9) | <0.001(32.25) |
| High | 231(9.6) | 79(34.2) | 152(65.8) |  |
| LDL-C (≥3.3mmol/l) |  |  |  |  |
| Normal | 2077(86.8) | 1083(52.1) | 994(47.9) | 0.858(0.03) |
| High | 317(13.2) | 167(52.7) | 150(47.3) |  |
| HDL-C (≤1.04mmol/l) |  |  |  |  |
| Normal | 2129(88.9) | 1142(53.6) | 987(46.4) | <0.001(15.68) |
| High | 317(13.2) | 108(40.8) | 157(59.2) |  |
| Triglyceride (≥1.7mmol/l) |  |  |  |  |
| Normal | 1862(77.8) | 1011(54.3) | 851(45.7) | <0.001(14.58) |
| High | 530(22.1) | 238(44.9) | 292(55.1) |  |
| Measures of adiposity |  |  |  |  |
| BMI (kg/m<sup>2</sup>) |  |  |  |  |
| Underweight | 38(1.6) | 21(55.3) | 17(44.7) | 0.066(7.2) |
| Normal | 1502(62.7) | 794(52.9) | 708(47.1) |  |
| Overweight | 612(25.6) | 329(53.8) | 283(46.2) |  |
| Obese | 240(10.0) | 106(44.2) | 134(55.8) |  |
| Waist circumference (≥94cm men, ≥80cm women) |  |  |  |  |
| Normal | 1882(78.6) | 870(46.2) | 1012(53.8) | 0.003(8.569) |
| Abnormal | 512(21.4) | 274(53.5) | 238(46.5) |  |
| WHtR risk |  |  |  |  |
| Normal (≤0.5) | 1050(43.9) | 36(3.4) | 1014(96.6) | <0.001(1786.73) |
| Increased risk (0.51- 0.59) | 1276(53.3) | 1146(89.8) | 130(10.2) |  |
| High risk (>0.6) | 68(2.8) | 68(100) | 0(0.0) |  |
| WHR (≥0.90 men, ≥0.85 women) |  |  |  |  |
| Low (≤0.5) | 763(31.9) | 753(98.7) | 10(1.3) | 0.210(3.12) |
| Moderate (0.51- 0.59) | 273(11.4) | 271(99.3) | 2(0.7) |  |
| High (≥0.6) | 207(8.6) | 207(100) | 0(0.0) |  |
| Target Organ Damage (TOD) |  |  |  |  |
| ECG-LVH (mm/mV) |  |  |  |  |
| NO | 2009(83.9) | 951(47.3) | 1058(52.7) | 0.315(1.010) |
| YES | 385(16.1) | 193(50.1) | 192(49.9) |  |
| LVMI (g/m<sup>2</sup>) |  |  |  |  |
| NO | 2055(85.8) | 969(47.2) | 1086(52.8) | 0.127(2.329) |
| YES | 339(14.2) | 175(51.6) | 164(48.4) |  |
| eGFR stages (mL/min/1.73 m2) |  |  |  |  |
| Stage I | 2123(88.7) | 1124(52.9) | 999(47.1) | 0.018(10.06) |
| Stage II | 169(7.1) | 77(45.6) | 92(54.4) |  |
| Stage III | 96(4.0) | 49(51.0) | 47(49.0) |  |
| Stage IV | 6(0.3) | 0(0.0) | 6(100) |  |
| eGFR (mL/min/1.73 m2) |  |  |  |  |
| Normal | 2120(88.6) | 1022(48.2) | 1098(51.8) | 0.251(1.318) |
| Abnormal | 274(11.4) | 122(44.5) | 152(55.5) |  |

**Table 2.** Mean (±SD) of specific demographic, clinical and biochemical characteristics of participants stratified by sex

| Characteristics | Total, Mean(±SD) | Female, Mean (±SD) | Male, Mean(±SD) | P-value |
| --- | --- | --- | --- | --- |
| Age (year) | 41.9(12.3) | 42(12) | 42(13) | 0.550 |
| BMI (kg/m<sup>2</sup>) | 24.8(4.7) | 24.62(4.4) | 25.04(4.99) | 0.029 |
| WC (cm) | 87.1(8.3) | 93.63(4.47) | 80.0(5.01) | <0.001 |
| WHtR | 0.5(0.05) | 0.54(0.03) | 0.46(0.03) | <0.001 |
| WHR | 0.88(0.05) | 0.94(0.06) | 0.81(0.06) | <0.001 |
| SBP (mmHg) | 127.8(23.3) | 128.9 (23.5) | 128.4(23.3) | 0.516 |
| DBP (mmHg) | 85.7(11.3) | 86(11) | 85(12) | 0.188 |
| Triglyceride | 1.65(0.34) | 1.63(0.32) | 1.66(0.35) | 0.013 |
| Total Cholesterol (mmol/l) | 4.97(0.72) | 4.89(0.66) | 5.05(0.77) | <0.001 |
| HDL-C (mmol/l) | 1.29(0.27) | 1.31(0.24) | 1.28(0.29) | 0.016 |
| LDL-C (mmol/l) | 2.99(0.66) | 3.00(0.65) | 2.98(0.66) | 0.315 |
| TC/HDL | 4.25(2.29) | 4.06(1.98) | 4.46(2.58) | <0.001 |
| LDL/HDL | 2.51(1.25) | 2.47(1.19) | 2.56(1.30) | 0.085 |
| Non-HDL-C | 3.67(0.84) | 3.59(0.75) | 3.77(0.92) | <0.001 |
| Non-HDL/HDL | 3.25(2.29) | 3.06(1.98) | 3.46(2.58) | <0.001 |
| FBS (mmol/l) | 5.10(1.64) | 5.16(1.67) | 5.04(1.61) | 0.080 |
| HbA1C (%) | 5.21(1.03) | 5.21(1.09) | 5.20(0.96) | 0.964 |
| Creatinine Level (ummol/l) | 79.63(21.19) | 86(22) | 73(17) | <0.001 |
| eGFR (mL/min/1.73 m2) | 98.37(15.63) | 100(15) | 97(16) | <0.001 |

### Cardiometabolic risk factors of study participants

As shown in Table 1, the prevalence of hypertension was 35.3%, diabetes mellitus was 8.3% and combined overweight and obesity (O/O) was 35.6%. In comparison with women, gender differences were not significant in Systolic Blood Pressure (128.9 ± 23.5mmHg versus 128.4 ± 23.3mmHg, p = 0.516) and Diastolic Blood Pressure (86.0 ± 11.2mmHg versus 85.0. ± 12.6mmHg, p =0.188). Anthropometric data also revealed significant gender differences in WHtR risk (p<0.001), WC (p<0.001), and WHR (p<0.001).

### Association between demographic characteristics and cardiovascular risk factors

The association between baseline demographic characteristics and cardiovascular risk factors were presented in Table 3. Multivariate logistic regression analysis for hypertension showed that age group (30-39 years) [OR = 0.163; 95% C.I: (0.079 - 0.336), p <0.001], high income > SLE 2,000 [OR = 0.574; 95% C.I.: (0.421, 0.782), p <0.001], unemployed [OR = 2.100; 95% C.I (1.407-3.134), p<0.001] and self-employed [OR = 1.912; 95% C.I: (1.282-2.849), p=0.001] were independently associated with hypertension. The odds ratio of having hypertension was strongest with unemployment. Diabetes mellitus shows a significant association with the age group 30-39 years [OR = 0.093; 95% C.I.: (0.023 - 0.378), p <0.001], income > SLE 2,000 [OR = 0.548; 95% C.I.: (0.348 - 0.865), p <0.001], income 1100-2000 [OR = 0.376; 95% C.I.: (0.239 - 0.591), p <0.001], and income 500-1,000 [OR = 0.376; 95% C.I.: (0.239 - 0.591), p <0.001]. Dyslipidemia was associated with the age group 40-49years [OR = 0.255; 95% C.I.: (0.121 - 0.537), p <0.001], and all occupational groups including self-employment [OR = 5.210; 95% C.I.: (3.123 - 8.691), p <0.001], unemployment [OR = 2.440; 95% C.I.: (1.469-4.052) p = 0.001], retired [OR = 2.085; 95% C.I.: (1.276 - 3.408), p = 0.003], student [OR = 4.389, 95% C.I. (1.778-10.834)]. Overweight/Obesity was significantly associated with all educational levels: primary education [OR = 5.781; 95% C.I. (4.181-7.994), p <0.001], secondary education [OR = 7.595, 95% C.I. (5.378-10.726), p<0.001], tertiary education [OR=2.220, 95% C.I (1.605-3.071), p >0.001] and unemployment [OR =0.647, 95% C.I. (0.452-0.925), p < 0.001]. For alcohol, the regression analysis shows an independent association in all age groups, and all educational levels, while smoking as a risk factor was only associated with participants earning SLE 1,100 – 2000. Waist circumference was associated with the age group 40 – 49 years and the various cadre of occupation.

**Table 3.** MULTIVARIABLE ASSOCIATIONS BETWEEN DEMOGRAPHIC CHARACTERISTICS AND CARDIOVASCULAR RISK FACTORS

|  |  |  |  |  |  |  |  |  |  |  |  |  |  |  |
| --- | --- | --- | --- | --- | --- | --- | --- | --- | --- | --- | --- | --- | --- | --- |
|  | HYPERTENSION |  | DIABETES |  | DYSLIPIDEMIA |  | OVERIGHT/OBESE |  | ALCOHOL |  | SMOKING |  | WAIST CIRCUM |  |
| VARIABLE | OR(95%CI) | P VALUE | OR(95%CI) | P VALUE | OR(95%CI) | P VALUE | OR(95%CI) | P VALUE | OR(95%CI) | P VALU E | OR(95%CI) | P VALUE | OR (95%CI) | P VALU E |
| Age, by group |  |  |  |  |  |  |  |  |  |  |  |  |  |  |
| 20-29 | Ref. |  | Ref. |  | Ref. |  | Ref. |  | Ref. |  | Ref. |  | Ref. |  |
| 30-39 | 0.163(0.079 - 0.336) | <0.001 | 0.093(0.023-0.378) | 0.001 | 0.548(0.256-1.174) | 0.122 | 0.648(0.312-1.348) | 0.246 | 0.051(0.022-0.115) | <0.001 | 2.598(0.339-19.940) | 0.358 | 0.548(0.256-1.174) | 0.122 |
| 40-49 | 0.493(0.248 - 0.978) | 0.043 | 0.677(0.224-2.045) | 0.489 | 0.255(0.121-0.537) | <0.001 | 0.785(0.384-1.603) | 0.506 | 0.064(0.029-0.142) | <0.001 | 3.149(0.418-23.748) | 0.266 | 0.255(0.121-0.537) | <0.001 |
| 50-59 | 0.581(0.292 - 1.155) | 0.121 | 0.802(0.263-2.440) | 0.697 | 0.851(0.410-1.770) | 0.666 | 0.655(0.319-1.342) | 1.342 | 0.052(0.023-0.115) | <0.001 | 2.346(0.309-17.817) | 0.410 | 0.851(0.410-1.770) | 0.666 |
| >60 | 0.959(0.478 -1.927) | 0.907 | 1.412(0.463-4.307) | 0.544 | 1.686(0.805-3,529) | 0.166 | 0.500(0.239-1.044) | 0.065 | 0.224(0.102-0.496) | <0.001 | 2.643(0.344-20.310) | 0.350 | 1.686(0.805-3.529) | 0.166 |
| Income |  |  |  |  |  |  |  |  |  |  |  |  |  |  |
| 0-500 | Ref. |  |  |  | Ref. |  | Ref. |  | Ref. |  | Ref. |  | Ref. |  |
| 500-1000 | 0.814(0.619 -1.070) | 0.140 | 0.392(0.258-0.596) | <0.001 | 0.762(0.551-1.055) | 0.101 | 0.989(0.740-1.322) | 0.940 | 0.958(0.658-1.393) | 0.821 | 0.729(0.473-1.122) | 0.151 | 0.762(0.551-1.055) | 0.101 |
| 1100-2000 | 0.613(0.458 -1.070) | 0.001 | 0.376(0.239-0.591) | <0.001 | 0.880(0.628-1.234) | 0.459 | 1.056(0.779-1.432) | 0.725 | 0.851(0.573-1.264) | 0.424 | 0.623(0.390-0.994) | 0.047 | 0.880(0.628-1.234) | 0.459 |
| >2000 | 0.574(0.421, 0.782) | <0.001 | 0.548(0.348-0.865) | <0.001 | 0.817(0.570-1.170) | 0.270 | 0.955(0.691-1.320) | 0.780 | 0.831(0.550-1.257) | 0.384 | 0.753(0.465-1.219) | 0.248 | 0.817(0.570-1.170) | 0.270 |
| Marital status |  |  |  |  |  |  |  |  |  |  |  |  |  |  |
| single | Ref. |  | Ref. |  | Ref. |  | Ref. |  | Ref. |  | Ref. |  | Ref. |  |
| Married | 1.030(0.668 -1.589) | 0.894 | 0.975(0.510-1.864) | 0.939 | 1.261(0.759-2.097) | 0.371 | 1.268(0.809-1.986) | 0.300 | 0.933(0.521-1.670) | 0.821 | 1.466(0.652-3.297) | 0.355 | 1.261(0.759-2.097) | 0.371 |
| Seperated/Divorce | 1.353(0.872 - 2.099) | 0.177 | 0.574(0.290-1.138) | 0.112 | 1.143(0.681-1.919) | 0.612 | 1.407(0.893-2.217) | 0.141 | 0.740(0.408-1.341) | 0.424 | 1.686(0.744-3.822) | 0.211 | 1.143(0.681-1.919) | 0.612 |
| Widow | 1.104(0.706 - 1.724) | 0.665 | 0.718(0.361-1.427) | 0.344 | 0.854(0.503-1.452) | 0.561 | 1.465(0.925-2.322) | 0.104 | 0.630(0.344-1.154) | 0.381 | 1.423(0.620-3.266) | 0.406 | 0.854(0.503-1.452) | 0.561 |
| Education level |  |  |  |  |  |  |  |  |  |  |  |  |  |  |
| None | Ref. |  | Ref. |  | Ref. |  | Ref. |  | Ref. |  | Ref. |  |  |  |
| Primary | 1.314(1.007 - 1,715) | 0.054 | 1.257(0.787-2.008) | 0.338 | 0.907(0.661-1.246) | 0.547 | 5.781(4.181-7.994) | <0.001 | 0.023(0.016-0.033) | <0.001 | 0.910(0.581-1.424) | 0.678 | 0.907(0.661-1.246) | 0.547 |
| Secondary | 0.972(0.721 - 1.312) | 0.855 | 1.271(0.754-2.141) | 0.368 | 0.809(0.565-1.156) | 0.244 | 7.595(5.378-10.726) | <0.001 | 0.016(0.010-0.026) | <0.001 | 1.043(0.638-1.706) | 0.866 | 0.809(0.565-1.156) | 0.244 |
| Tertiary | 0.981(0.760 - 1.265) | 0.882 | 1.328(0.856-2.062) | 0.205 | 1.065(0.793-1.430) | 0.674 | 2.220(1.605-3.071) | <0.001 | 0.065(0.048-0.087) | <0.001 | 0.941(0.619-1.431) | 0.777 | 1.065(0.793-1.430) | 0.674 |
| Occupation |  |  |  |  |  |  |  |  |  |  |  |  |  |  |
| Employed | Ref. |  | Ref. |  | Ref. |  | Ref. |  | Ref. |  | Ref. |  | Ref. |  |
| Self employed | 1.912(1.282 - 2.849) | 0.001 | 1.182(0.619-2.255) | 0.612 | 5.210(3.123-8.691) | <0.001 | 0.794(0.552-1.144) | 0.216 | 1.601(0.997-2.572) | 0.051 | 0.828(0.452-1.516) | 0.540 | 5.210(3.123-8.691) | <0.001 |
| Unemployed | 2.100(1.407 - 3.134) | <0.001 | 0.714(0.355-1.434) | 0.344 | 2.440(1.469-4.052) | 0.001 | 0.647(0.452-0.925) | 0.017 | 1.163(0.727-1.862) | 0.529 | 0.842(0.465-1.523) | 0.569 | 2.440(1.469-4.052) | 0.001 |
| Retired | 1.616(1.113 - 2.345) | 0.012 | 0.626(0.335-1.169) | 0.142 | 2.085(1.276-3.408) | 0.003 | 0.860(0.620-1.194) | 0.368 | 0.971(0.628-1.502) | 0.894 | 0.986(0.577-1.684) | 0.959 | 2.085(1.276-3.408) | 0.003 |
| Student | 1.320(0.584 - 2.984) | 0.504 | 1.719(0.476-6.203) | 0.408 | 4.389(1.778-10.834) | 0.001 | 0.435(0.189-1.004) | 0.051 | 0.144(0.056-0.371) | <0.001 | 4.885(0.592-40.301) | 0.141 | 4.389(1.778-10.83) | 0.001 |
| Religion |  |  |  |  |  |  |  |  |  |  |  |  |  |  |
| Christian | Ref. |  | Ref. |  | Ref. |  | Ref. |  | Ref. |  | Ref. |  | Ref. |  |
| Muslim | 1.092(0.901 - 1.323) |  | 1.020(0.732-1.420) | 0.908 | 1.227(0.979-1.538) | 0.076 | 1.430(1.175-1.740) | 0.000 | 1.053(0.813-1.362) | 0.697 | 0.917(0.661-1.272) | 0.604 | 1.227(0.979-1.538) | 0.076 |

### Preclinical Target Organ Damage (TOD) of the study participants

In this study, 16.1% had ECG-Left ventricular hypertrophy, while 14.2% had an abnormal Left Ventricular Wall Mass Index (LVMI) by 2D Echo measurement. The participants’ impaired kidney function (eGFR) was 11.4%, with eGFR stage II being the highest at 7.1%. Men had a significantly higher risk of eGFR staging than women (p<0.018).

### Association of cardiometabolic risk factors with preclinical Tissue Organ Damage

Tables 4,5, & 6 show the multivariate binary logistic regression analysis results between CMFRs and the indices of target organ damage. The odds ratio of Cardiometabolic risk factor in relation to Target Organ Damage is also shown in Supplementary Figure 1.

**Table 4.** Association Between Cardiometabolic Risk Factors and Specific Target Organ Damage (ECG-LVH)

| ECG- LVH |  |  |  |  |  |  |
| --- | --- | --- | --- | --- | --- | --- |
| Variable | Total No.(%) | YES No.(%) | NO No.(%) | OR (95% CI) | Model 1 | Model 2 |
| **Overweight/OBESE** |  |  |  |  |  |  |
| NO | 1598(66.8) | 239(15.0) | 1359(85.0) | Ref. |  |  |
| YES | 796(33.2) | 146(18.3) | 650(81.7) | 0.793(0.631- 0.996) | 0.784(0.625-0.984) | 0.693(0.542 - 0.885) |
| **Alcohol** |  |  |  |  |  |  |
| NO | 1486(62.1) | 236(15.9) | 1250(84.1) | Ref. |  |  |
| YES | 908(37.9) | 149(16.4) | 759(83.6) | 0.938(0.748-1.176) | 0.880(0.688-1.125) | 1.032(0.756 -1.409) |
| **SMOKING** |  |  |  |  |  |  |
| NO | 2073(86.6) | 328(15.8) | 1745(84.2) | Ref. |  |  |
| YES | 321(13.4) | 57(17.8) | 264(82.2) | 0.871(0.637 - 1.176) | 0.751(0.518 -1.088) | 0.764(0.526 - 1.109) |
| **DIABETES** |  |  |  |  |  |  |
| NO | 2187(91.7) | 356(16.3) | 1831(83.7) | Ref. |  |  |
| YES | 199(8.3) | 27(13.6) | 172(86.4) | 1.255(0.822 - 1.916) | 1.276(0.833 - 1.955) | 1.325(0.861- 2.035) |
| **HYPERTENSION** |  |  |  |  |  |  |
| NO | 1550(64.7) | 237(15.3) | 1313(84.7) | Ref. |  |  |
| YES | 844(35.3) | 148(17.5) | 696(82.5) | 0.843(0.671, 1.058) | 0.859(0.680 - 1.084) | 0.856(0.676 -1.084) |
| **WAIST CIRCUMFERENCE** |  |  |  |  |  |  |
| NORMAL | 1882(78.6) | 300(15.9) | 1582(84.1) | Ref. |  |  |
| ABNORMAL | 512(21.4) | 85(16.6) | 427(83.4) | 0.952(0.730 - 1.240) | 0.968(0.736 - 1.273 | 0.948(0.717 - 1.254) |
| **TOTAL CHOLESTEROL** |  |  |  |  |  |  |
| NORMAL | 2163(90.4) | 344(15.9) | 1819(84.1) | Ref. |  |  |
| ABNORMAL | 231(9.6) | 41(17.7) | 190(82.3) | 0.742(0.454 - 1.210) | 0.901(0.624 - 1.302) | 0.860(0.593 -1.247) |
| **LDL-C** |  |  |  |  |  |  |
| NORMAL | 2077(86.8) | 327(15.7) | 1750(84.3) | Ref. |  |  |
| HIGH | 317(13.2) | 58(18.3) | 259(81.7) | 0.647(0.379 - 1.107) | 0.834(0.610 -1.142) | 0.811(0.586 -1.122) |
| **LOW HDL-C** |  |  |  |  |  |  |
| NO | 1985(82.9) | 311(15.7) | 1674(84.3) | Ref. |  |  |
| YES | 409(17.1) | 74(18.1) | 335(81.9) | 0.835(0.630 - 1.107) | 0.841(0.636 - 1.113) | 0.852(0.643 - 1.129) |
| **Dyslipidemia** |  |  |  |  |  |  |
| NO | 1882(78.6) | 300(15.9) | 1582(84.9) | Ref. |  |  |
| YES | 512(21.4) | 85(16.6) | 427(83.4) | 1.449(0.834 - 2.518) | 0.968(0.736 -1.273) | 0.948(0.717 - 1.254) |

**Table 5.** Association Between Cardiometabolic Risk Factors and Specific Target Organ Damage (LVMI)

| LVMI |  |  |  |  |  |  |
| --- | --- | --- | --- | --- | --- | --- |
| Variable | Total No.(%) | YES No.(%) | NO No.(%) | OR (95% CI) | Model 1 | Model 2 |
| **Overweight/OBESE** |  |  |  |  |  |  |
| NO | 1598(66.8) | 210(13.1) | 1388(86.9) | Ref. |  |  |
| YES | 796(33.2) | 129(16.2) | 667(83.8) | 0.790(0.621 - 1.005) | 0.777(0.612-0.987) | 0.712(0.540-0.921) |
| **Alcohol** |  |  |  |  |  |  |
| NO | 1486(62.1) | 208(14.0) | 1278(86.0) | Ref. |  |  |
| YES | 908(37.9) | 131(14.0) | 777(85.5) | 0.945(0.745-1.199) | 0.915(0.705-1.186) | 1.088(0.781 -1.516) |
| **SMOKING** |  |  |  |  |  |  |
| NO | 2073(86.6) | 289(13.9) | 1784(86.1) | Ref. |  |  |
| YES | 321(13.4) | 50(15.6) | 271(84.4) | 0.883(0.634 - 1.229) | 0.654(0.448-0.953) | 0.668(0.457-0.977) |
| **DIABETES** |  |  |  |  |  |  |
| NO | 2187(91.7) | 313(14.3) | 1874(85.7) | Ref. |  |  |
| YES | 199(8.3) | 25(12.6) | 174(87.4) | 1.176(0.759 - 1.823) | 1.197(0.770-1.863) | 1.259(0.805-1.970) |
| **HYPERTENSION** |  |  |  |  |  |  |
| NO | 1550(64.7) | 209(13.5) | 1341(86.5) | Ref. |  |  |
| YES | 844(35.3) | 130(15.4) | 714(84.4) | 0.857(0.674 -1.088) | 0.869(0.680-1.111) | 0.867(0.676-1.112) |
| **WAIST CIRCUMFERENCE** |  |  |  |  |  |  |
| NORMAL | 1882(78.6) | 265(14.1) | 1617(85.9) | Ref. |  |  |
| ABNORMAL | 512(21.4) | 74(14.5) | 194(84.0) | 0.956(0.724 - 1.264) | 1.017(0.762-1.358) | 1.010(0.751-1.357) |
| **TOTAL CHOLESTEROL** |  |  |  |  |  |  |
| NORMAL | 2163(90.4) | 302(14.0) | 1791(86.02) | Ref. |  |  |
| ABNORMAL | 231(9.6) | 37(16.0) | 264(83.3) | 0.646(0.387 - 1.079) | 0.908(0.619-1.332) | 0.861(0.584 - 1.269) |
| **LDL-C** |  |  |  |  |  |  |
| NORMAL | 2077(86.8) | 286(13.8) | 1791(86.3) | Ref. |  |  |
| HIGH | 317(13.2) | 53(16.7) | 264(83.3) | 0.501(0.280 - 0.894) | 0.810(0.585-1.123) | 0.821(0.585-1.152) |
| **LOW HDL-C** |  |  |  |  |  |  |
| NO | 1985(82.9) | 271(13.7) | 1714(86.3) | Ref. |  |  |
| YES | 409(17.1) | 68(16.6) | 341(83.4) | 0.768(0.573 - 1.028) | 0.789(0.590-1.056) | 0.801(0.597-1.073) |
| **Dyslipidemia** |  |  |  |  |  |  |
| NO | 1882(78.6) | 265(14.1) | 1617(85.9) | Ref. |  |  |
| YES | 512(21.4) | 74(14.5) | 438(85.5) | 1.844(1.006 - 3.380) | 1.017(0.762-1.358) | 1.010(0.751 -1.357) |

**Figure 1:**
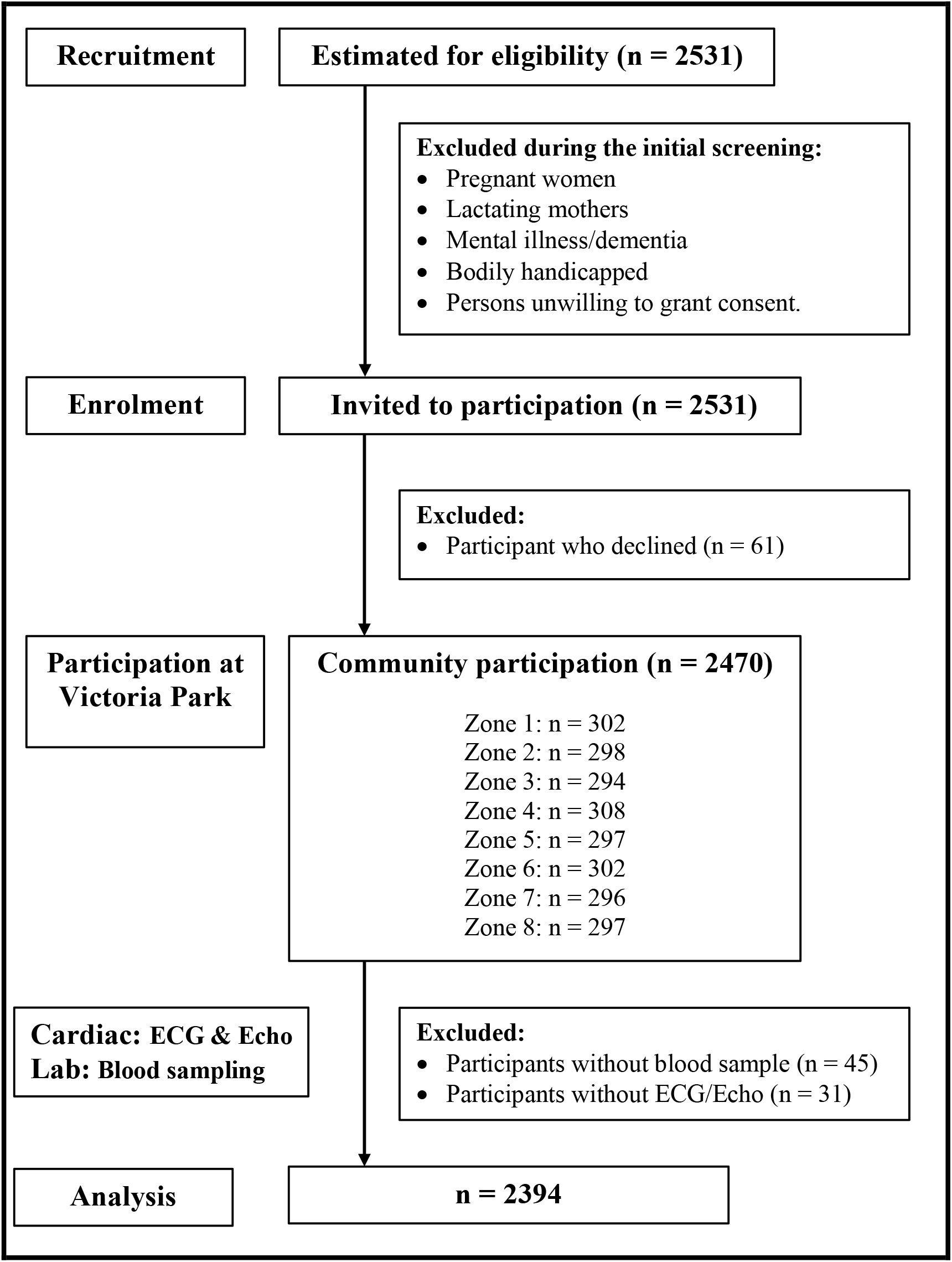
Steps involved during recruitment of participants and final analysis of data.

In table 4, diabetes mellitus [OR = 1.176, 95% C.I. (0.759 - 1.823)] and dyslipidaemia [OR = 1.844, 95% C.I. (1.006 -3.380)] were strongly associated with LVH. After adjusting sex and age for potential confounders, model 1 showed that DM was the only CMRF associated with LVH, while in model 2, alcohol and diabetes were associated with LVH. Table 5 shows that the multivariate binary logistic regression analysis for LVMI shows a strong association with diabetes mellitus [OR =1.176, 95% C.I (0.759-1.823)] and dyslipidemia [OR = 1.844, 95% C.I (1.006-3.380)]. Upon adjusting age and sex to determine the influence of potential confounders, Model 1 analysis established an association of diabetes mellitus, dyslipidemia, and waist circumference with LVMI, while Model 2 showed an additional cofounder of alcohol being associated with LVMI. For CKD (Table 6), the multivariate analysis shows that the odds of having CKD was strongly associated with diabetes mellitus [OR =1.212, 95% CI (0.741-1.983)], hypertension [OR =1.163, 95% CI (0.887-1.525)], alcohol [OR =1.003, 95% CI (0.772, 1.303)], Low HDL-C [OR = 1.261, 95% CI (0.881, 1.804)], High LDH-C [OR=1.355 95% CI (0.754, 2.433)] and TC [OR =1.170, 95% CI (0.663-2.066)]. Regression (model 1 & model 2) adjustment analysis demonstrated diabetes mellitus and high low HDL-C as the strongest determinant for LVH.

**Table 6.** Association Between Cardiometabolic Risk Factors and Specific Target Organ Damage (CKD)

| CKD |  |  |  |  |  |  |
| --- | --- | --- | --- | --- | --- | --- |
| Variable | Total No.(%) | YES No.(%) | NO No.(%) | OR (95% CI) | Model 1 | Model 2 |
| **Overweight/OBESE** |  |  |  |  |  |  |
| NO | 1598(66.8) | 168(10.5) | 1430(89.5) | Ref. |  |  |
| YES | 796(33.2) | 106(13.3) | 690(86.7) | 0.750(0.578, 0.974) | 0.774(0.596 - 1.005) | 0.778(0.589 -1.028) |
| **Alcohol** |  |  |  |  |  |  |
| NO | 1486(62.1) | 172(11.6) | 1314(88.4) | Ref. |  |  |
| YES | 908(37.9) | 102(11.2) | 1844(89.0) | 1.003(0.772, 1.303) | 0.915(0.688 -1.218) | 0.810(0.565 -1.161) |
| **SMOKING** |  |  |  |  |  |  |
| NO | 2073(86.6) | 229(11.0) | 1844(89.0) | Ref. |  |  |
| YES | 321(13.4) | 45(14.0) | 276(86.0) | 0.784(0.554, 1.111) | 0.678(0.448-1.024) | 0.665(0.439-1.008) |
| **DIABETES** |  |  |  |  |  |  |
| NO | 2187(91.7) | 254(11.6) | 1933(88.4) | Ref. |  |  |
| YES | 199(8.3) | 19(9.5) | 180(90.5) | 1.212(0.741, 1.983) | 1.200(0.730-1.975) | 1.208(0.730 -1.997) |
| **HYPERTENSION** |  |  |  |  |  |  |
| NO | 1550(64.7) | 186(12.0) | 1364(88.0) | Ref. |  |  |
| YES | 844(35.3) | 88(10.4) | 756(89.6) | 1.163(0.887 - 1.525) | 1.126(0.852-1.486) | 1.118(0.843 - 1.483) |
| **WAIST CIRCUMFERENCE** |  |  |  |  |  |  |
| NORMAL | 1882(78.6) | 213(11.3) | 1669(88.7) | Ref. |  |  |
| ABNORMAL | 512(21.4) | 61(11.9) | 451(88.1) | 0.926(0.683 - 1.254) | 0.870(0.634 -1.193) | 0.825(0.598 - 1.139) |
| **TOTAL CHOLESTEROL** |  |  |  |  |  |  |
| NORMAL | 2163(90.4) | 247(11.4) | 1916(88.6) | Ref. |  |  |
| ABNORMAL | 231(9.6) | 27(11.7) | 204(88.3) | 1.170(0.663 - 2.066) | 0.876(0.566 - 1.356) | 0.862(0.555 - 1.339) |
| **LDL-C** |  |  |  |  |  |  |
| NORMAL | 2077(86.8) | 239(11.5) | 1838(88.5) | Ref. |  |  |
| HIGH | 317(13.2) | 35(11.0) | 282(89.2) | 1.355(0.754 - 2.433) | 1.000(0.682 -1.465) | 0.951(0.641 -1.409) |
| **LOW HDL-C** |  |  |  |  |  |  |
| NO | 1986(82.9) | 235(11.8) | 1751(88.2) | Ref. |  |  |
| YES | 409(17.1) | 39(9.5) | 370(90.5) | 1.261(0.881 - 1.804) | 1.277(0.893 -1.827) | 1.275(0.890 - 1.825) |
| **Dyslipidemia** |  |  |  |  |  |  |
| NO | 1882(78.6) | 213(11.3) | 1669(88.7) | Ref. |  |  |
| YES | 512(21.4) | 61(11.9) | 451(88.1) | 0.706(0.399 - 1.252) | 0.870(0.634 - 1.193) | 0.825(0.598 - 1.139) |

The relationship between clinical sensitivity and specificity for ECG-LVH as a target organ damage by gender was evaluated using the Receiver Operating Characteristic (ROC) curve. A low optimal cut-off point for ECG-LVH (male 24.5 vs female 27.5mm) was required to maximize sensitivity and specificity (Fig. 2). Additional information on the sensitivity and specificity of parameters related to target organ damage is shown in Supplementary Table 1.

**Figure 2.**
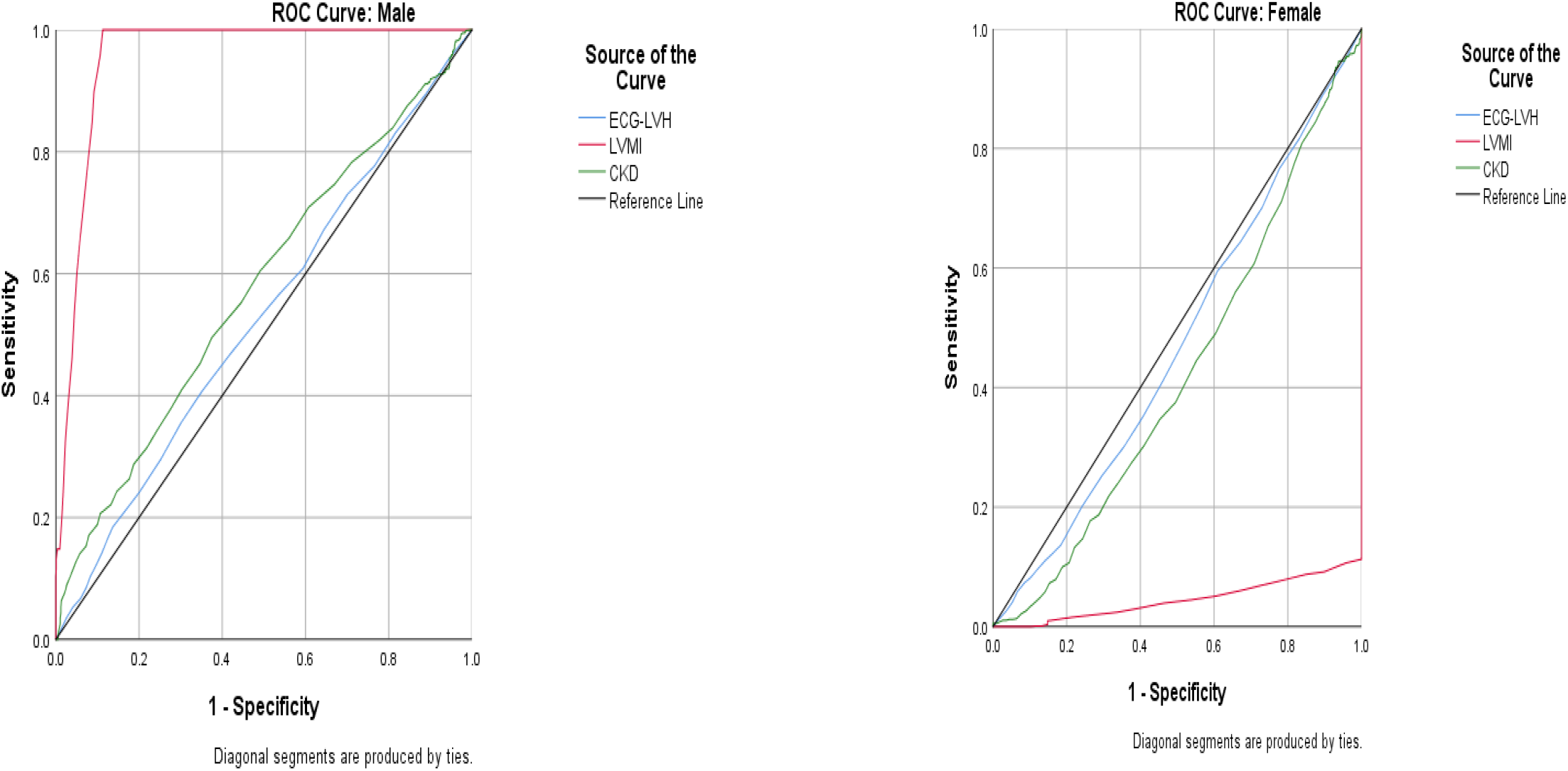
Area under the curve for specific TOD (LVH, LVMI, eGFR)

## DISCUSSION

Health screenings are essential for identifying cardiovascular disease risk and its attending mortality and morbidity. These cardiometabolic risk factors are necessary to predict its burden and complications (kidney disease, stroke and coronary artery disease) and may adversely influence the quality of life of the individual [38]. This notwithstanding, population-based screening remains limited in LMICs. Our study provides the largest data on CMRFs in health screening for NCD in Sierra Leone.

It is the first study to characterize the distribution of cardiometabolic risk factors and preclinical target organ damage among adults in Sierra Leone. Our findings indicate that CMRFs are common among adult Sierra Leoneans with hypertension, diabetes and dyslipidemia having the strongest association with specific preclinical TOD. The study suggests a high prevalence of cardiometabolic risk factors for CVD, as many Sierra Leonean adults have at least one significant risk factor: hypertension (35.6%), diabetes mellitus (8.3%), Overweight/Obesity (37.3%), abdominal obesity (21.4%), dyslipidaemia (21.4%), and alcohol consumption (37.7%). The reported prevalence in our study is consistent with findings from other studies in SSA [39-41]. In Sierra Leone, the observed patterns of CMRFs indicate that a demographic health transition might be occurring faster than previously reported [22,24,26]. Therefore, our study has contributed critical evidence on the burden and distribution of CMRFs among adults living in an urban setting in SSA.

This study’s prevalence of hypertension (35.5%) is similar to other community-based studies in SSA [42-44]. This prevalence of hypertension was identical to the previous WHO STEPS survey in 2009 that reported 37% in males and 33% in females [22]. The study design and age population of 25-65 years used in the STEPs survey make it difficult to compare with our study. In Sierra Leone, a much higher prevalence of hypertension (49.6%) was recently reported in a provincial district by Odland et al., while a lower rate of hypertension (22%) was reported by Geraedts et al when compared to this study [24,26]. The disparity may be attributed to the age differences of the studied cohorts (20 years and above in our study, unlike 40 years and above in the reported study by Orland et al. [24]). The difference may also be ascribed to the study design, socio-demographic characters, and lifestyle patterns of the study participants. Our estimated prevalence (8.3%) of type 2 diabetes mellitus is higher than the prevalence reported from other studies in Sierra Leone – 3.5% in 2009, 5.5% in 2021, 6.2% in 2017, 2.4% in the urban population and 0% in the rural population in 1997 [23,24,45]. The high urban prevalence of diabetes in our study was partly due to the combined use of FBG and HBA1C, unlike other studies conducted in Sierra Leone and the greater variance in fasting glucose among urban participants. Additionally, the high prevalence of DM in this study compared to previous studies could be partly attributable to previous studies being 10 -15 years earlier. Even though the prevalence of diabetes is higher in this study, the small population size and methodology used in previous studies would make comparisons difficult.

Overweight (26.5%) and obesity (10.0%) were surprisingly more common in our study, as Sierra Leone is one of the poorest countries in the world. The estimated 36.5% of overweight/obese reported in this study is higher than the 25% reported by Orland and colleagues, the first study to evaluate CVDRFs in a larger sample size in Sierra Leone [24]. Our study’s estimated finding of O/O is consistent with other studies from Ghana, Nigeria, and Ethiopia. [46,47,48]. Our study’s high proportion of individuals with increased BMI may suggest an upward trend in this risk factor, thereby supporting the hypothesis of rapid urbanization and a westernized lifestyle. Previous studies have indicated that waist circumference, as an indicator of abdominal obesity, correlates positively with a risk for cardiovascular diseases [49]. Abdominal obesity was more common in our study, with men more likely than females to be affected, probably because of the tendency of central obesity. Similarly, a study conducted in Ethiopia showed waist circumference to be associated with hypertension [50].

Even though BMI is an independent cardiometabolic risk for cardiovascular diseases, there is evidence strongly suggesting Waist-Height-Ratio (WHtR) > 0.5 as the highest predictor of all cardiometabolic risk factors for both sexes, even more than BMI and WC combined [51,52]. When WHtR was analyzed, more than half of the study participants were categorized into *“increased risk (53*.*3)”* and *“high-risk (2*.*8)”*. WHtR as a predictor of cardiovascular events was generally higher in our study than BMI and WC. This result confirmed earlier findings in existing literature [52]. The results of WHtR >0.5 allow us to conclude that more adults Sierra Leoneans are at “early health risk” for cardiovascular disease.

The previous perception that dyslipidaemia was rare among Black Africans is now being discredited by several studies showing a high prevalence of dyslipidaemia among Black Africans [53,54]. In this study, elevated TG (22.1%) was the most prominent form of dyslipidemia, followed by an elevated LDL-C and HDL-C, with women having the highest prevalence of all measures of dyslipidaemia in comparison to men. This observed pattern of dyslipidaemia prevalence in our study is similar to a survey conducted in Ghana [55] but inconsistent with results documented by Asiki and team [56] and Gebreegziabiher et al [57], where the most prevalent dyslipidaemia markers were HDL-C, TC and LDL-C. Despite the observed disparities in the different measures of dyslipidaemia, studies have reported a high prevalence of all forms of dyslipidaemia among women [58-61]. This study further demonstrated that women were more likely to have high levels of low HDL-C albeit the widely accepted belief that HDL is male-specific [56,57]. These findings illustrate the importance of health screening for dyslipidaemia as a large proportion of the study participants is dyslipidemic.

WHO has identified several major risk factors for cardiovascular disorder, including smoking, alcohol consumption, unhealthy diets and physical inactivity [3,31]. About one-third of the participants had consumed alcohol in this study, and the rate/frequency of consumption was high. Our report is higher than the WHO-reported general prevalence of alcohol consumption in most SSA countries [3]. The increased consumption of alcohol in our study could partly be attributed to our youthful participants, that comprised about half of the cohort with the ability to afford its cost. It was observed in this study that one-third of participants do not engage in any form of exercise, with women being less educated, unemployed, and physically inactive than their men counterparts. These results are consistent with reports from a Ghanaian study [55]. Cigarette smoking was generally uncommon in our study, but the impact of “Shesha pipe smoking” among young age must be evaluated.

Our analysis to identify the association between CMRFs and some demographic variables revealed that hypertension was associated with the youthful age group, non-employment status and increased income, while diabetes mellitus was associated with youthful and increased income. Dyslipidaemia was associated with middle age and non-employment. Education level, all age groups and being a student were associated with alcohol consumption. Earning more income was associated with smoking, while young age and all employment status were associated with waist circumference. Our study’s findings are consistent with several SSA studies [4,12,62,63] and confirm our earlier statement that cardiometabolic risk factors are the principal causes of cardiovascular diseases in Sierra Leone.

We investigate the role of CMRFs in developing preclinical target organ damage (TOD). Studies on Left Ventricular Hypertrophy are scarce in Africa because of the non-availability of electrocardiograms and echocardiograms in many settings. However, few studies on the Black population living in Africa show an overall prevalence of LVH of 4.1% in Ghana, 62% in Cameroon, 41% in the Gambia, and 41% in Angola [55, 64-66]. In our study, the prevalence of LVH by ECG and LVMI were 16.1% and 12.4%, respectively. Our findings were higher than the Ghanaian study [65] but comparatively lower than other African reports. The odds of having LVH either by ECG or LVMI were further evaluated in this study, and our findings demonstrated a strong association with diabetes and dyslipidaemia. Other studies have found hypertension to have a strong association with LVH, even though it is inconsistent with our findings. The weak association of hypertension with LVH in this study could be attributed to our youthful study population (about half of the population is less than age 40 years), as most of the hypertensives were young. Studies have reported that LVH in hypertensives is increased several-fold with ageing and in hypertensives with risk factor-adjusted cardiovascular morbidity, which was unlike our study [67,68]

Using the regression model adjustment analysis, diabetes mellitus was identified as the strongest determinant for LVH in our study. Other studies have reported LVH to be common among diabetic patients, with LVH being a strong predictor of cardiovascular disease in diabetics [69,70]. Since hypertension is a low predictor of LVH in this study, a ROC curve was performed to show the relationship between clinical sensitivity and specificity for ECG - LVH cut-off. This demonstrated that a low cut-off points for ECG-LVH (male 24.5mm vs female 27.5mm) were required to maximize sensitivity and specificity. This analysis suggests that LVH may occur at a much lower cut-off for Sierra Leoneans and that the standard cut-off points for LVH may fail as a screening tool for target organ damage in this setting. These findings need further research and in-depth evaluation in future studies. The prevalence of CKD in our population was 11.6%, and the odds of having CKD was strongly associated with diabetes mellitus, hypertension, alcohol, Low HDL-C, High LDH-C and TC. Regression (model 1 & model 2) adjustment analysis demonstrated diabetes mellitus and high low HDL-C as the strongest determinant for LVH. These findings confirmed the recent results by Coker et al who reported diabetes mellitus as the second most common cause of CKD for admission into a tertiary hospital in Sierra Leone, while Chakimanga and colleagues reported a high prevalence of 29.9% CKD in Rural Sierra Leone [71,72]. Hence the strong association of diabetes mellitus as a risk factor for CKD observed in this study is a wake-up call for action on kidney disease screening and prevention programs in Sierra Leone.

Our findings should be interpreted within the context of the following limitation. Since the study is cross-sectional in design, it could not conclude direct causality inference of risk factors and effect outcomes. Additionally, as a health screening study, some of the clinical outcomes were not repeated, and this may result in measurement errors, with the potential of underestimating cardiometabolic risk factors. CKD assessment by single serum creatinine without assessing for proteinuria, which indicates the presence of CKD, will also lead to an underestimation of CKD. However, the findings in our study are consistent with other large prospective studies in developing countries.

## Conclusion

The study provides novel data-driven information on the burden of cardiometabolic risk and its association with target organ damage, as it is the first health screening survey on a larger population in Sierra Leone. This study’s relatively high prevalence of cardiometabolic risk factors indicates that CVD is increasing in Sierra Leone, a country whose health services are already overburdened by tuberculosis, malaria, and HIV/AIDS. Despite the various assumptions underlying these projections, the importance of this work cannot be overestimated. The result of this study could serve as the basis for advocacy with an urgent call for action in the establishment of programmes that would improve the control and management of cardiometabolic risk factors and CVD, along with other NCDs.

## Data Availability

All data produced in the present study are available upon reasonable request to the authors

https://www.researchregistry.com/browse-the-registry#home/

## Twitter

James B.W. Russell@AProfJamesBWRu,

## Ethics approval and registration

The research proposal, questionnaire and consent form were approved by the Sierra Leone Ethics and Scientific Review Committee. The institution does not provide ethics reference identification number. Anonymity was maintained using serial coded numbers assigned to the case records and the extracted data was handled with strict confidentiality. The protocol of this study also registered under Research Registry with the unique identification number researchregistry8201, that is available at https://www.researchregistry.com/browse-the-registry#home/

## Acknowledgements

The authors acknowledge the financial support provided by Ecobank Sierra Leone Limited. We thank Dr Jattu Rahman-Sesay, Dr. Tejan Mansaray and Dr. Jajuah for coordinating the screening campaign at Victoria Park. We express gratitude to the 4^th^ year medical students (2019) and nurses who served as data collectors including the field manager who worked tirelessly in ensuring that this study was successful. These medical students have graduated as Medical Doctors and they include: Dr Mohamed Samura, Dr Abdul Karim Bah, Dr Evelyn Hawa Kamara, Dr Chernor Abubakarr Barrie, Paul Thoronka, Dr Osman Kanneh, Dr Alieu Kanu, Dr Vidal Dupigny, Dr Scholastica Nduisi, Dr Omar Bah. A big thank to nurses from the Ministry of Health and Sanitation: Zainab Kargbo, Fatmata Koroma, Abigail Pratt, Claudia Campbell, Angel Jones, Lovetta Davies, Fatmata Bangura, Albert Rogers, Patrick Coker, Abibatu Jones, Silvia Kanyako and Gillian Jones. We are also grateful for the technical laboratory assistance provided by the Ecomed Advance Medical Laboratory. Additionally, we thank the staff of Prime Care Medical Clinic: Fatmata Nicol, Belinda Mattia, Mariatu Turay, and George Russell who supported in the cardiac screening of the participants. We wish to thank all the participants enrolled in this study.

## Author’s Contributions

**JWBR**, conceptualized, designed the overall study, formal analysis and writing – original draft manuscript. **SS** conceptualized, and design the overall study, **TRK**, data curation, formal analysis writing and writing – original draft manuscript. **SKS**, data curation, and formal statistical analysis. **SL, OTA, AB, JSK, DRL**, reviewing and editing. **VC, MS**, project administration and community recruitment of participants. All authors substantively reviewed the manuscripts inputted into revisions and approved the final manuscript.

## Patient consent for publication

Not required.

## Provenance and peer review

Not commissioned, externally peer-reviewed.

## Data availability statement

The anonymised dataset supporting this study’s findings is available upon reasonable request from the corresponding author as cited in the publication. Access for further investigation and analysis will be granted to researchers following a methodologically sound proposal submitted after publication.

## Abbreviations

BMI: Body Mass Index
CMD: Cardiometabolic Disease
CMRF: Cardiometabolic risk factors
CVD: cardiovascular disease
CVDRF: cardiovascular disease risk factors
DBP: diastolic blood pressure
LDL-C: Low density lipoprotein cholesterol
MIC: low-and middle-income countries
LVH: left ventricular hypertrophy
LVMI: Left ventricular mass index
NCD: Non-communicable disease
HDL-C: High density lipoprotein cholesterol
SBP: systolic blood pressure
SLE: Leones currency
TG: triglyceride
TOD: target organ damage
WC: waist circumference
WHO: World Health Organization

## Figure legend

**Supplementary Figure 1**. Odds ratio of Cardiometabolic risk factor in relation to Target Organ Damage Dys = Dyslipidemia, LHDL-C = Low High-Density Lipoprotein, LDL-C Low = Density Lipoprotein, TC = Triglyceride, WC = Waist Circumference, Hp =Hypertension Dia = Diabetes Mellitus

